# Elevated plasma neurofilament light & glial fibrillary acidic protein in epilepsy versus non-epileptic seizures & non-epileptic disorders

**DOI:** 10.1101/2024.02.19.24303018

**Authors:** Hannah Dobson, Said Al Maawali, Charles Malpas, Alexander F Santillo, Matthew Kang, Marian Todaro, Rosie Watson, Nawaf Yassi, Kaj Blennow, Henrik Zetterberg, Emma Foster, Andrew Neal, Dennis Velakoulis, Terence John O’Brien, Dhamidhu Eratne, Patrick Kwan

**Author notes:** Corresponding Author: Prof Patrick Kwan.

## Abstract

**Background:** Research suggests that recurrent seizures may lead to neuronal injury. Neurofilament light chain protein (NfL) and glial fibrillary acidic protein (GFAP) levels increase in cerebrospinal fluid and blood following neuroaxonal damage, and have been hypothesised as potential biomarkers for epilepsy. We examined plasma NfL and GFAP levels and their diagnostic utility in differentiating patients with epilepsy from those with psychogenic non-epileptic seizures (PNES), and other non-epileptic disorders.

**Methods:** We recruited consecutive adults admitted for video-electroencephalography monitoring and formal neuropsychiatric assessment. Plasma samples were collected on admission. NfL and GFAP levels were quantified and compared between patient groups and an age-matched reference cohort (n=1,926), and correlated with clinical variables.

**Results:** 149 patients were included. 115 were diagnosed with epilepsy, 22 with PNES and 12 with other conditions. Plasma NfL and GFAP levels were elevated in patients with epilepsy compared to PNES, adjusted for age and sex (NfL p=0.004, GFAP p=0.004). A significantly higher proportion of patients with epilepsy (26%) had NfL levels above the 95^th^ age-matched percentile compared to the reference cohort (5%; p=0.0265). NfL levels above the 95^th^ percentile of the reference cohort had a 97% positive predictive value for epilepsy.

**Discussion:** Elevated NfL or GFAP levels may support an underlying epilepsy diagnosis and caution against a diagnosis of PNES alone. Further examination of associations between NfL and GFAP levels and specific epilepsy subtypes or seizure characteristics may provide valuable insights into disease heterogeneity and contribute to the refinement of diagnosis, understanding pathophysiological mechanisms, and formulating treatment approaches.

## INTRODUCTION

Epilepsy is diagnosed when a person experiences more than one unprovoked epileptic seizure in a period greater than 24 hours, or evidence of high risk of recurrence after a single seizure(1). Diagnosis is based on patient and witness reports, careful review of a patient’s medical history including descriptions of their seizures, investigation findings, particularly neuroimaging and electroencephalography (EEG)(2–4).

In contrast to epileptic seizures psychogenic non-epileptic seizures (PNES) were not accompanied by epileptiform activity in the EEG before, during or after the seizure(5). Owing to their resemblance in behavioural manifestation, in the absence of video-EEG recorded clinical events, PNES may be misdiagnosed as epileptic seizures(6). Delayed recognition of psychogenic non-epileptic seizures is associated with increased psychosocial morbidity(7,8), as well as increased mortality risk(9), highlighting the importance of an early, accurate diagnosis. In addition, an incorrect diagnosis of epilepsy may impact a person’s quality of life and have significant economic consequences due to stigma, lifestyle constraints and adverse effects of anti-seizure medications (ASM) and missing non-epilepsy diagnoses(2,10).

There is an increasing body of research examining the use of blood biomarkers in the diagnosis and differential diagnoses of a range of neurological and neurodegenerative disorders(11–13). Recent research has identified that biomarkers such as neurofilament light chain protein (NfL), a marker of neuronal injury, can be used to distinguish neurological and neurodegenerative disorders, from primary psychiatric and non-neurodegenerative disorders(13–16).

Clinical observation, neuropsychological assessments and neuroimaging studies suggest that recurrent seizures may lead to neuronal injury in some patients(17,18). NfL and glial fibrillary acidic protein (GFAP) are neuronal cytoplasmic proteins which are highly expressed in myelinated axons and astrocytes, respectively. NfL and GFAP levels increase in cerebrospinal fluid and blood in response to neuroaxonal damage(11,12), and they have been hypothesised as potential biomarkers for epilepsy(19). Studies have reported higher NfL levels in adult patients admitted with status epilepticus compared to those with chronic epilepsy(20,21), and in post-stroke epilepsy(22) and autoimmune epilepsy compared with chronic epilepsy and PNES(23).

NfL has been reported to be elevated in patients with epilepsy compared with controls in some studies(20,24). GFAP has been shown to be increased in adults and children with epilepsy compared with controls(25,26) and adult patients with non-epileptic seizures in patients admitted to an EMU(27). Whilst promising, these studies did not focus on distinguishing epilepsy from important differential diagnoses such as PNES and included relatively small patient populations.

The primary aim of this study was to quantify plasma NfL and GFAP levels and their diagnostic utility in differentiating patients with epilepsy from those with PNES, and other non-epileptic disorders. We included patients who had undergone ‘gold-standard’ diagnosis with video EEG monitoring, multidisciplinary assessments, and investigations, with the final diagnosis determined at a multidisciplinary case conference. We hypothesised that NfL and GFAP levels would be higher in patients with epilepsy compared to those with non-epileptic disorders. We further compared NfL levels against a large reference cohort to examine for any associations between NfL and clinical variables in patients with epilepsy(16,28). A similar reference cohort is not available for GFAP levels.

## METHODS

### Study design and participants

We prospectively recruited consecutive patients at least 18 years of age admitted to the Epilepsy Monitoring Unit (EMU) at The Alfred Hospital in Melbourne, Australia, between June 2018 and July 2022. The study, part of The Markers in Neuropsychiatric Disorders Study (The MiND Study, https://themindstudy.org), was approved by Human Research Ethics Committees at the Alfred Hospital (157/19 and 611/20), and The Royal Melbourne Hospital (MH/HREC2020.142). All patients provided written informed consent.

Patients were admitted to the EMU for video-EEG monitoring for diagnostic clarification, classification of seizure type and optimisation of epilepsy treatment including consideration of surgical interventions. Detailed demographic and clinical data were obtained, including age and sex, age of seizure onset, seizure frequency over the preceding 12 months, and seizure frequency. Seizure frequency was classified according to the Seizure Frequency Scoring System(29). Neuroimaging was performed or reviewed. All patients were reviewed by a board-certified neuropsychiatrist.

At the conclusion of the EMU admission, results were reviewed by a multidisciplinary team which comprised epileptologists, neuroradiologists, neuropsychiatrists, neuropsychologists, and neurophysiologists. Patients were diagnosed with epilepsy, psychogenic non-epileptic seizures (PNES), or other conditions based upon the comprehensive multidisciplinary review. In this analysis, patients diagnosed to have both epileptic and non-epileptic seizures were included in the epilepsy category. The medical record was reviewed for follow-up information, and the most up to date diagnosis was used for this study.

### Laboratory procedure and NfL and GFAP measurements

On EMU admission, plasma samples were collected in a purple-topped ethylenediaminetetraacetic acid (EDTA) tube, centrifuged (2,000 revolutions per minute) for ten to fifteen minutes, and stored at −80 degrees Celsius at the Alfred Neuroscience Bio-Databank, a biorepository and databank dedicated to research in neurological disorders(30). Plasma aliquots were analysed for NfL and GFAP using the Quanterix Simoa HD-X platform at the Walter and Eliza Hall Institute, Melbourne, Australia.

### Statistical analyses

Statistical analyses were performed using R v4.2.2 (2022-10-31). General linear models (GLMs) were used to examine relationships between NfL and GFAP, and diagnostic group, and relevant clinicodemographic variables. The dependent variables were log_10_-transformed biomarker levels; diagnostic group, age, and sex were independent variables. 95% confidence intervals were computed (nonparametric bootstrapping, 1000 replicates), with statistical significance defined as any confidence interval not including the null hypothesis value (at 95% level). These statistical methods were selected because they mitigate the effects of distributional violations, including presence of outliers. Receiver operator characteristic (ROC) curves were computed to estimate area under the curve (AUC) for distinguishing between groups. The optimal cut-off was determined using Youden’s method and relevant classification metrics were then computed (sensitivity, specificity, positive predictive value, negative predictive value, positive likelihood ratio, and negative likelihood ratio).

NfL levels from all patient cohorts were compared to a large reference control cohort. This cohort and models and methodology have been described in detail previously(16,28). Briefly, this cohort included 1,926 people aged 5–90 years, with no history or clinical symptoms or signs of neurological disorder, with plasma NfL analysed using the Quanterix Simoa HD-X platform. Z-scores were calculated from this reference cohort, derived using generalised additive models for location, scale, and shape (GAMLSS). Single-sample t-tests were used to test the null hypothesis that the mean z-score was 0 (i.e., no difference/equal to the mean of reference control group). Welch’s independent samples t-tests were used to compare z-scores between groups. Where appropriate, *p* values were computed with a critical alpha level 0.05 used to define statistical significance.

## RESULTS

During the study period, 324 patients were admitted to the Alfred EMU, of whom 152 consented to participate in this study. Three patients were excluded from analysis because their detailed medical records were unavailable. The final cohort of 149 patients included 115 patients diagnosed with epilepsy, including 3 with both epilepsy and PNES. 22 had psychogenic non-epileptic seizures, and 12 other conditions (‘Other’). The Other group included cardiac causes (n=5), migraines (n=3), non-diagnostic non-epilepsy (n=1), provoked seizure (alcohol) (n=1), dementia (n=1), and idiopathic primary hypersomnolence (n=1),

The mean ages of patients in the Epilepsy, PNES, and Other groups were 40.8 years (standard deviation [SD] 15.1), 38.5 years (SD 11.8), and 48 years (SD 21.3), respectively. There was a higher proportion of females in the PNES group (90.9%), compared to 54.8% in epilepsy, and 75% in Other. Further details are available in Tables 1 and 2.

**Table 1:** Demographic data.

|  | **Epilepsy** | **PNES** | **Other** |
| --- | --- | --- | --- |
|  | ***N=115*** | ***N=22*** | ***N=12*** |
| Age (years) | 40.8 (15.1) | 38.5 (11.8) | 48.0 (21.3) |
| Sex: Female | 63 (54.8%) | 20 (90.9%) | 9 (75.0%) |
| NfL (pg/mL) | 12.4 (16.5) | 6.53 (3.23) | 14.5 (17.7) |
| Log10NfL | 0.93 (0.33) | 0.77 (0.20) | 0.95 (0.43) |
| NfL z-score<br>(age<br>adjusted) | 0.76 (1.36) | 0.22 (0.98) | 0.49 (1.23) |
| GFAP | 107 (116) | 61.4 (28.2) | 107 (75.0) |
| Log10GFAP | 1.90 (0.30) | 1.74 (0.20) | 1.93 (0.31) |

GFAP: glial fibrillary acidic protein; NfL: neurofilament light chain protein
The Other group included: cardiac causes (n=5), migraines (n=3), non-diagnostic non-epilepsy (n=1), provoked seizure (alcohol) (n=1), dementia (n=1), idiopathic primary hypersomnolence (n=1)
GFAP: glial fibrillary acidic protein; NfL: neurofilament light chain protein; PNES: psychogenic non-epileptic seizures.

**Table 2:** Characteristics of epilepsy cohort in patients with epilepsy with elevated NfL levels (>95^th^ percentile compared to controls), and not elevated levels.

|  | [ALL epilepsy] | <=95th<br>percentile | >95th<br>percentile | p.overall | N |
| --- | --- | --- | --- | --- | --- |
|  | *N=115* | *N=85* | *N=30* |  |  |
| Age | 40.8 (15.1) | 43.0 (16.0) | 34.7 (10.1) | 0.002 | 115 |
| Sex: |  |  |  | 0.978 | 115 |
| Female | 63 (54.8%) | 46 (54.1%) | 17 (56.7%) |  |  |
| Male | 52 (45.2%) | 39 (45.9%) | 13 (43.3%) |  |  |
| NfL | 12.4 (16.5) | 7.56 (5.29) | 26.2 (26.8) | 0.001 | 115 |
| Log10NfL | 0.93 (0.33) | 0.81 (0.23) | 1.27 (0.34) | <0.001 | 115 |
| NfL z-score (age<br>adjusted) | 0.76 (1.36) | 0.13 (0.87) | 2.54 (0.80) | <0.001 | 115 |
| NfL percentile | 0.66 (0.31) | 0.54 (0.28) | 0.98 (0.02) | <0.001 | 115 |
| Localisation: |  |  |  | 0.434 | 115 |
| FLE | 15 (13.0%) | 10 (11.8%) | 5 (16.7%) |  |  |
| IGE | 9 (7.83%) | 7 (8.24%) | 2 (6.67%) |  |  |
| TLE | 49 (42.6%) | 40 (47.1%) | 9 (30.0%) |  |  |
| Unclassified | 37 (32.2%) | 24 (28.2%) | 13 (43.3%) |  |  |
| Unknown | 5 (4.35%) | 4 (4.71%) | 1 (3.33%) |  |  |
| Age of onset | 19.9 (17.4) | 21.4 (18.9) | 15.9 (11.8) | 0.071 | 114 |
| Seizure duration<br>(years) | 21.1 (17.5) | 21.9 (18.5) | 18.8 (14.3) | 0.349 | 115 |
| Seizure frequency(29) | 7.22 (1.97) | 7.15 (1.98) | 7.41 (1.97) | 0.569 | 99 |
| Seizure type |  |  |  | 0.794 | 102 |
| Focal | 69 (67.6%) | 49 (66.2%) | 20 (71.4%) |  |  |
| FTBTC | 17 (16.7%) | 12 (16.2%) | 5 (17.9%) |  |  |
| Generalised | 16 (15.7%) | 13 (17.6%) | 3 (10.7%) |  |  |
| MRI lesion |  |  |  | 0.700 | 89 |
| No | 53 (59.6%) | 40 (61.5%) | 13 (54.2%) |  |  |
| Yes | 36 (40.4%) | 25 (38.5%) | 11 (45.8%) |  |  |
| MRI lesion type |  |  |  | 0.881 | 36 |
|  | *N=115* | *N=85* | *N=30* |  |  |
| Cavernoma/vascular malformation | 4 (11.1%) | 3 (12.0%) | 1 (9.09%) |  |  |
| Gliosis/stroke | 2 (5.56%) | 2 (8.00%) | 0 (0.00%) |  |  |
| Hippocampal sclerosis | 8 (22.2%) | 5 (20.0%) | 3 (27.3%) |  |  |
| Hippocampal sclerosis + vascular | 2 (5.56%) | 1 (4.00%) | 1 (9.09%) |  |  |
| MCD/FCD | 9 (25.0%) | 5 (20.0%) | 4 (36.4%) |  |  |
| Other | 2 (5.56%) | 2 (8.00%) | 0 (0.00%) |  |  |
| Tumour/glioma | 9 (25.0%) | 7 (28.0%) | 2 (18.2%) |  |  |

GFAP: glial fibrillary acidic protein; NfL: neurofilament light chain protein; FLE: frontal lobe epilepsy; IGE: idiopathic generalised epilepsy; TLE: temporal lobe epilepsy; FTBTC: focal to bilateral tonic clonic; MCD: malformation of cortical development; FCD: focal cortical dysplasia

### Plasma NfL and GFAP in diagnostic groups

As demonstrated in Table 1 and Figure 1, NfL levels were higher in in patients with epilepsy compared to PNES. This difference was statistically significant and persisted after adjusting for age and sex (ß=0.41 [0.13, 0.67] p=0.004). GFAP levels were also higher in patients with epilepsy compared to patients with PNES (Figure 2). This difference was also significant after adjusting for age and sex (ß=0.49 [0.15, 0.80] p=0.004). NfL and GFAP levels were not different in the Other group, compared epilepsy and PNES groups.

**Figure 1.**
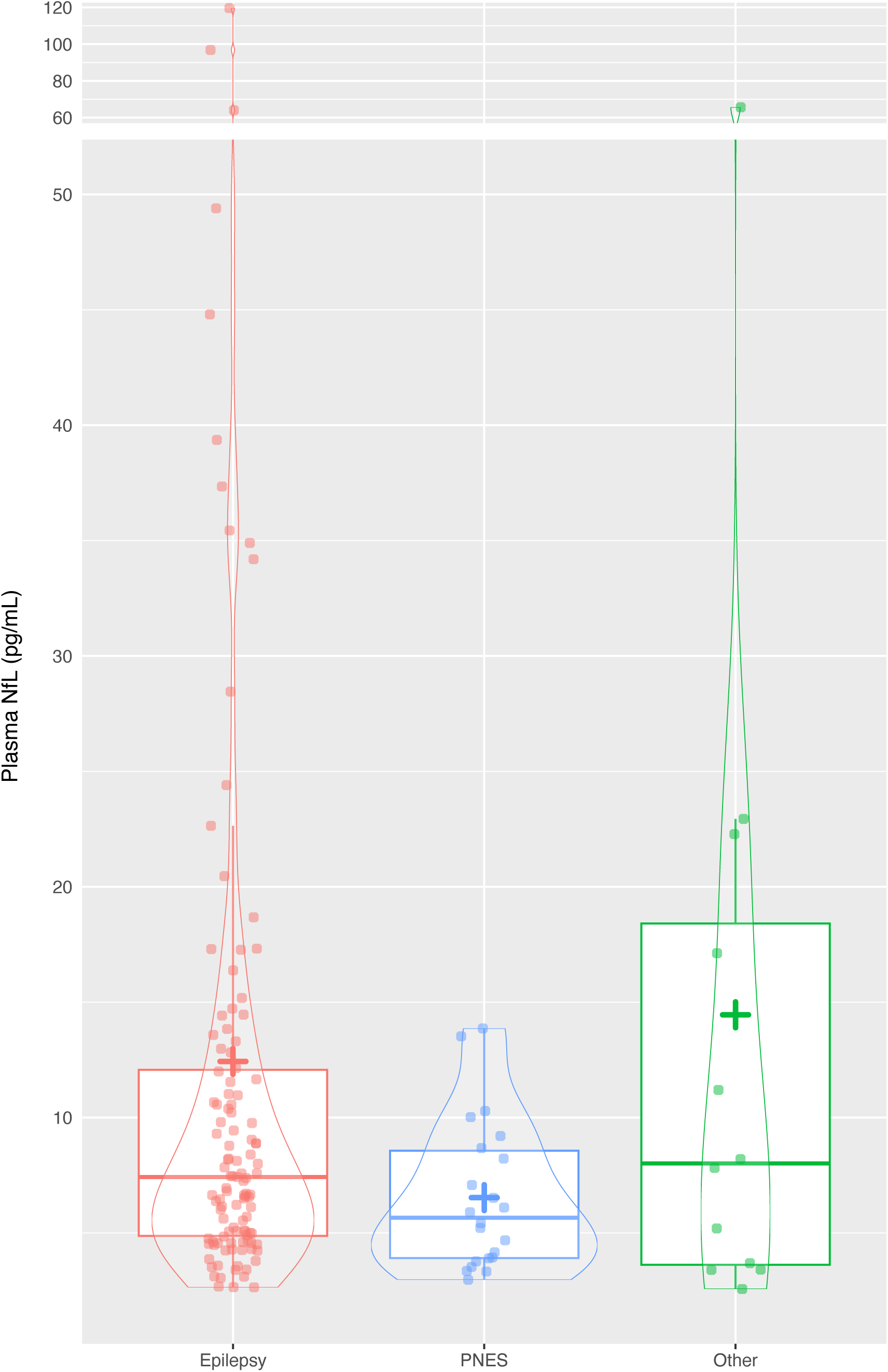
Plasma NfL levels in patients with epilepsy, psychogenic non-epileptic seizures, or other diagnoses. **+ = mean level**

**Figure 2.**
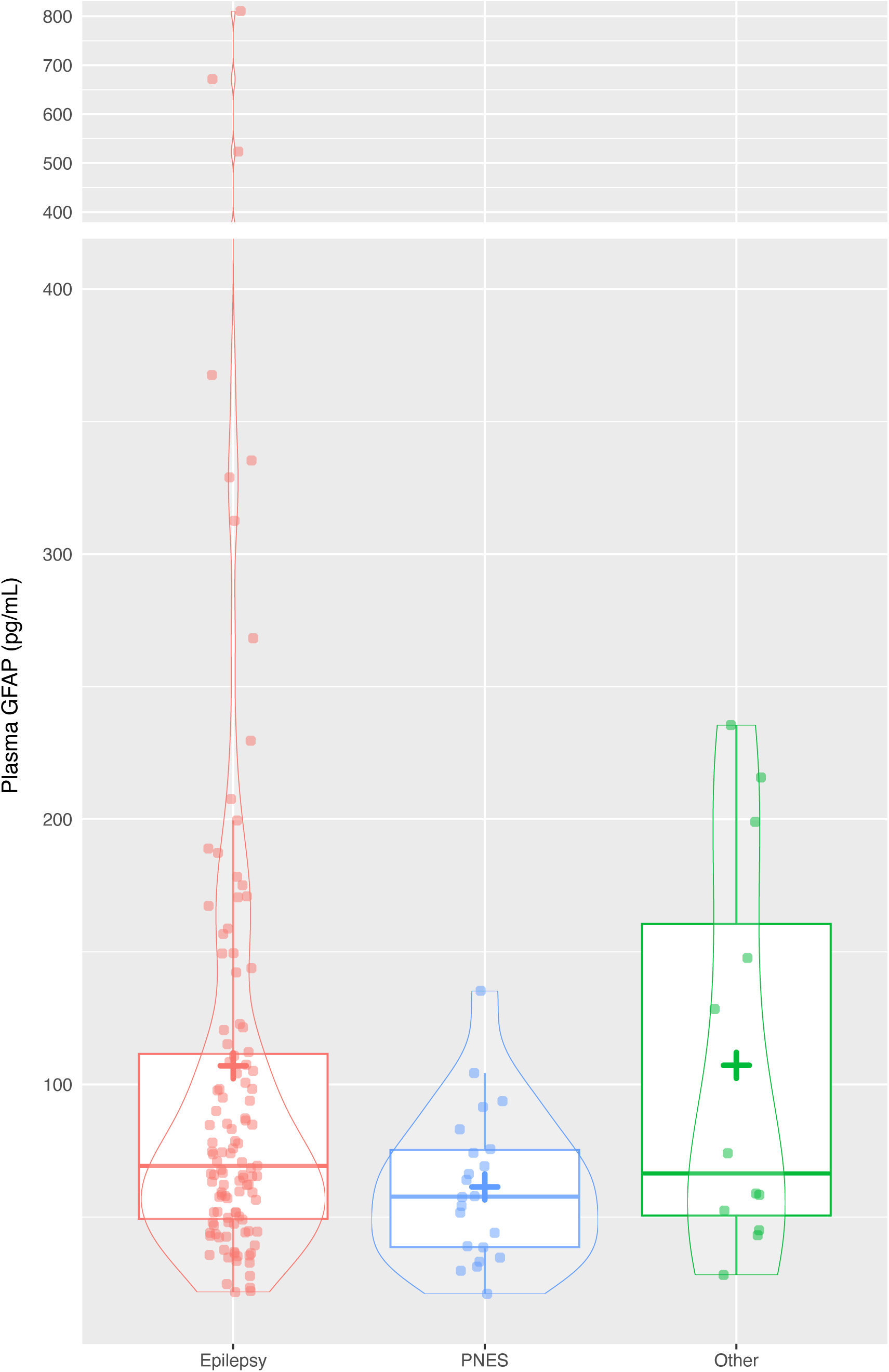
Plasma GFAP levels in patients with epilepsy, psychogenic non-epileptic seizures, or other disorders. **+ = mean levels**

### Diagnostic performance of plasma NfL and GFAP to distinguish epilepsy from PNES

Absolute plasma NfL level demonstrated weak ability to distinguish epilepsy from PNES (area under the curve, AUC=0.65 [0.52, 0.77]), Figure 3. Youden’s method revealed an optimal cut-off of 4.2pg/mL, which resulted in 36% specificity, 89% sensitivity. Looking at alternative cut-offs which optimised for specificity, a cut-off of 10.3pg/mL resulted in 91% specificity, 33% sensitivity. GFAP also demonstrated weak diagnostic performance (AUC 0.65 [0.53, 0.77]), with a cut-off of 93.8pg/mL associated with 91% specificity, 35% sensitivity, and an alternative cut-off optimising for sensitivity, 39.1pg/mL: 32% specificity, 87% sensitivity. There was no difference seen between performance of the two biomarkers, and combinations of biomarkers did not result in improved diagnostic performance.

**Figure 3.**
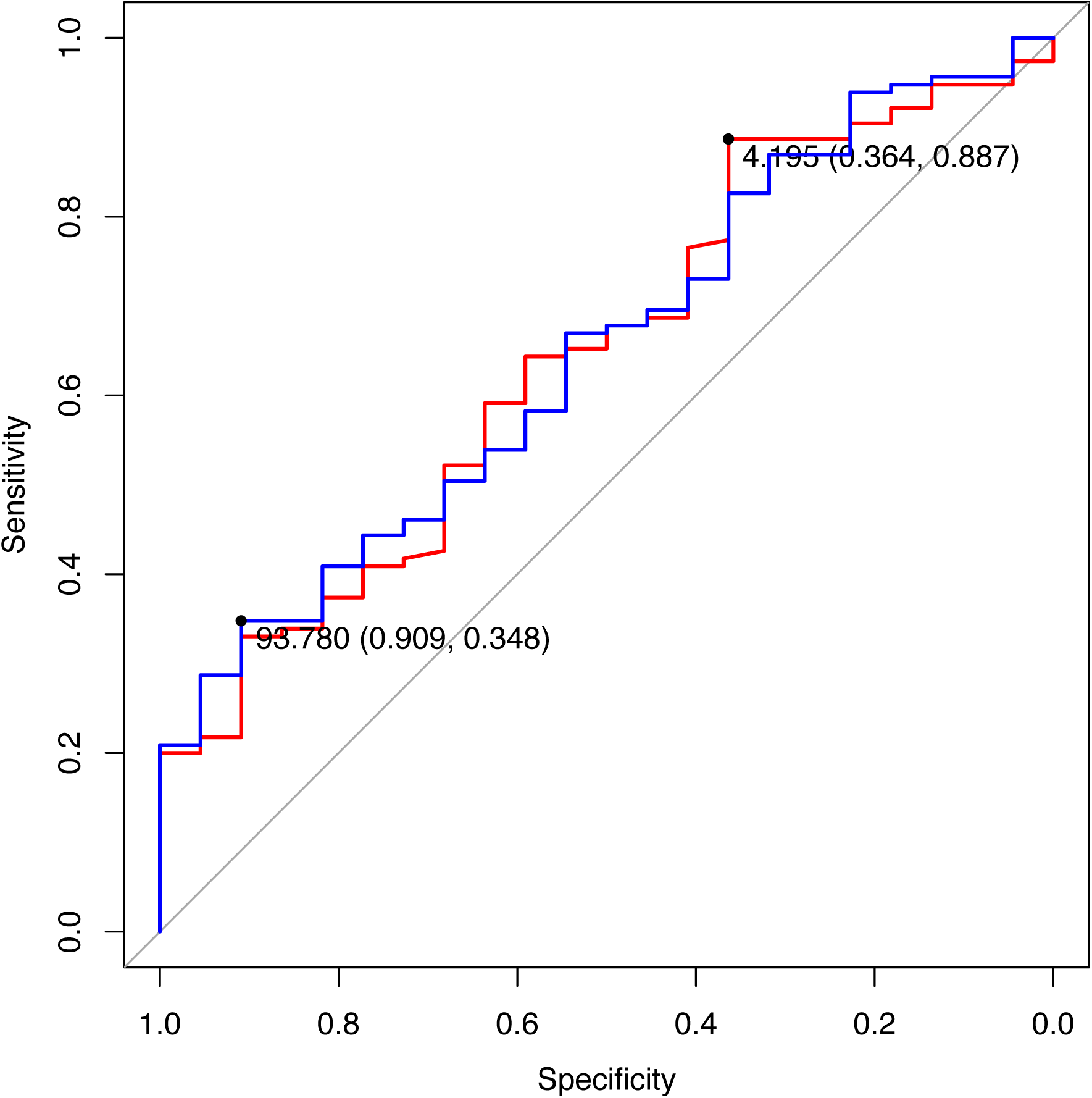
Receiver operator characteristic (ROC) curves of plasma NfL and GFAP levels to distinguish between patients with epilepsy and PNES. Red: NfL. Blue: GFAP. Numbers = area under the curve (specificity, sensitivity)

### Plasma NfL compared to a large reference control cohort

We compared age-adjusted z-scores derived from a large reference control cohort with NfL levels (Table 2 and Figure 5). This revealed significantly higher NfL levels in epilepsy compared to PNES (mean z-score 0.76 vs 0.22). Performing a GLM with NfL z-score as the dependent variable, and diagnostic group as independent variable, again showed higher levels in epilepsy compared to PNES (ß=0.41 [0.08, 0.76] p=0.006). Higher levels were also seen in epilepsy compared to the control group (0.76 vs 0, difference = 0.76, 95% CI [0.50, 1.01], p < .001). No evidence of difference was seen in PNES vs controls (p=0.315), epilepsy vs Other (p=0.495), and Other vs controls (p=0.196).

As demonstrated in Figure 4, a significantly greater than expected number/proportion of patients with epilepsy had NfL levels greater than the 95^th^ percentile for their age (30/114, 26%), compared to only 1/22 (5%) in PNES (p=0.0265). Notably, these were all in people younger than 60 years of age. Using the 95^th^ percentile as a cut-off, high NfL level was associated with 26% sensitivity, 95% specificity, 5.7 positive likelihood ratio, 0.77 negative likelihood ratio, 97% positive predictive value, 20% negative predictive value for a diagnosis of epilepsy. In the Other group, 2/12 (17%) had levels greater than the 95^th^ percentile. These patients were diagnosed with dementia and an alcohol-related acquired brain injury.

**Figure 4.**
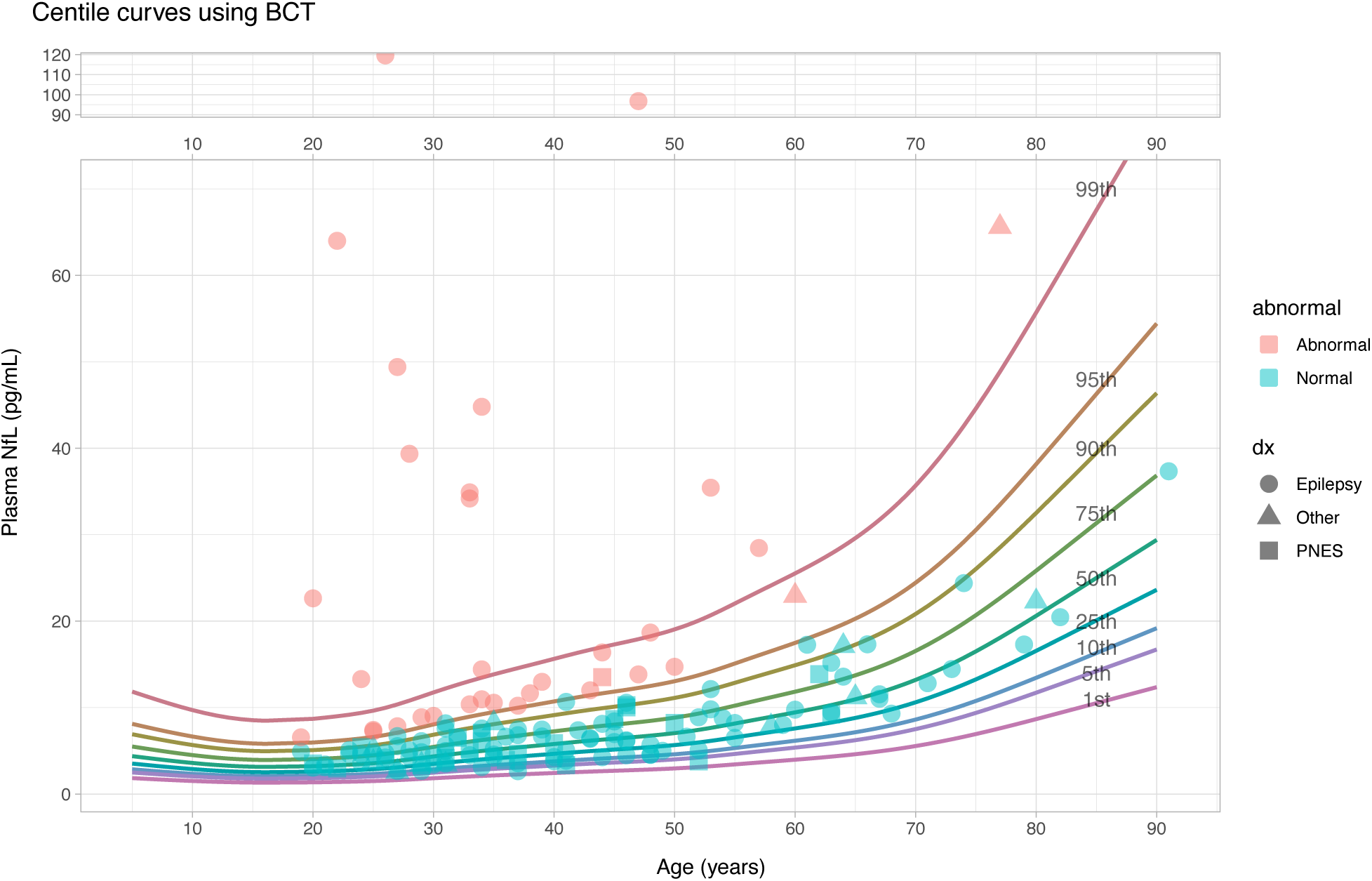
Plasma NfL levels in patents with epilepsy, PNES or other diagnoses compared to a large reference cohort. Levels >95^th^ percentile were defined as abnormal and those below 95^th^ percentile as normal.

**Figure 5.**
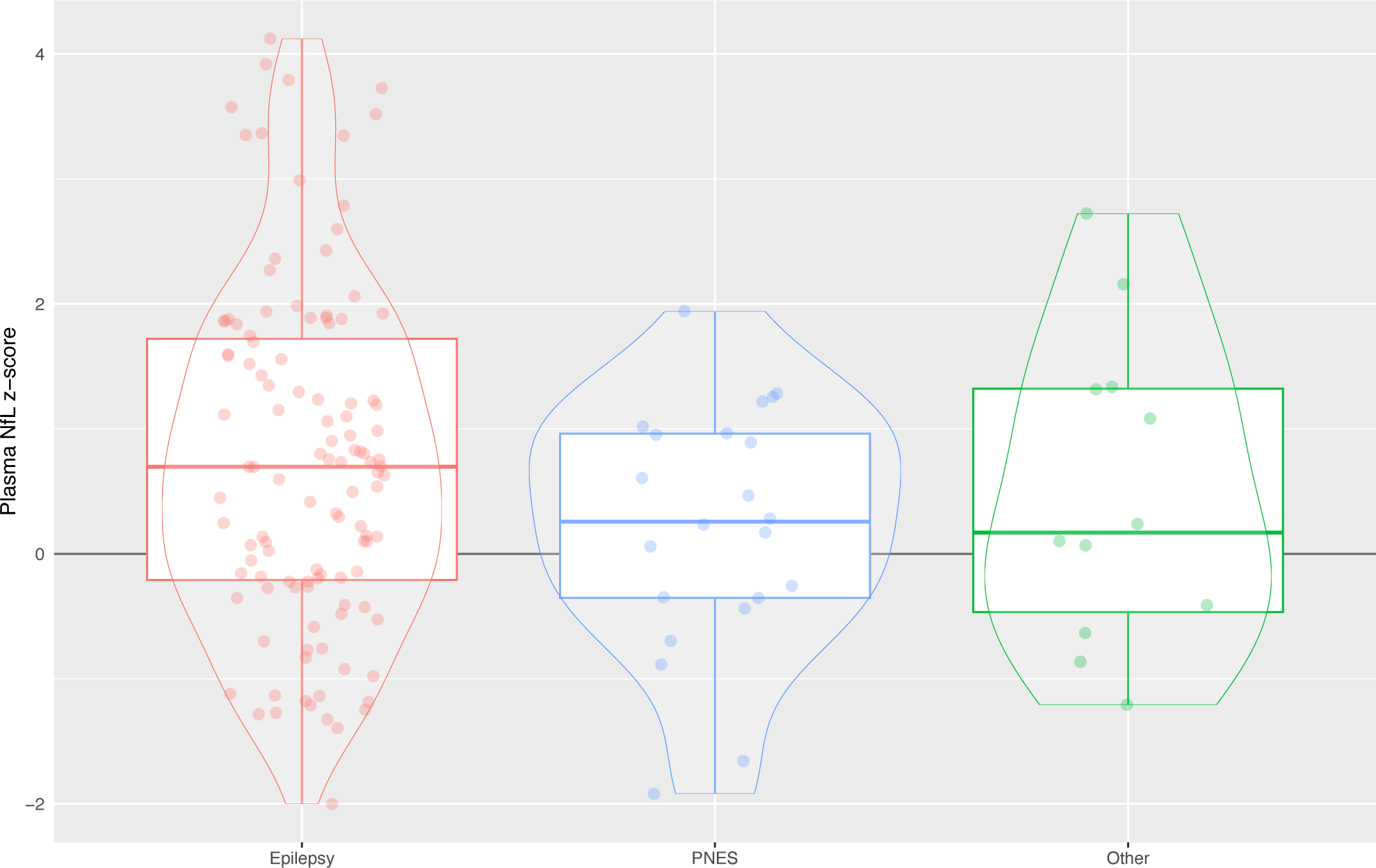
NfL level z-scores (age-adjusted, compared to large reference cohort)

### Biomarkers and clinical variables

Subgroup analysis was performed examining differences in clinical variables for patients with plasma NfL levels above the 95^th^ percentile, compared to those with plasma NfL levels less than or equal to the 95^th^ percentile (Table 2). Patients with plasma NfL levels above the 95^th^ centile were younger than those with lower plasma NfL levels (34.7 vs. 43 years, p=0.002). Proportions of lesional epilepsy, seizure frequency or seizure type, and a range of other clinical variables (see Table 2), were not different in the >95^th^ percentile compared to <=95^th^ percentile groups.

Given the lack of a large reference cohort for GFAP, we stratified GFAP levels in the epilepsy group into quartiles to explore differences in clinical variables between quartiles (Supplementary Material). There were no differences observed in clinical variables between quartiles of GFAP levels. Similarly, no differences were seen when performing the same analyses by quartiles of NfL levels.

## DISCUSSION

This study compared plasma NfL and GFAP levels in patients with epilepsy to those with PNES and other non-epileptic disorders, whose diagnoses were confirmed by video-EEG monitoring and formal neuropsychiatric evaluation. We identified elevated NfL and GFAP levels in patients with epilepsy compared to PNES and controls. NfL levels above the 95^th^ age-matched percentile of a large reference cohort were highly predictive of epilepsy rather than PNES, particularly in the younger adult population.

Few studies have compared NfL and GFAP levels between people with epilepsy, PNES and other non-epileptic disorders. Our findings suggest that patients with chronic epilepsy may experience some degree of ongoing neuronal injury. This observation seemed to be particularly pertinent in the younger adults (less than 60 years of age), for whom a highly elevated NfL (e.g., >95^th^ percentile compared to age-matched controls), and/or highly elevated GFAP (no patients with PNES had a GFAP level >150pg/mL), was highly predictive of epilepsy instead of PNES. This suggests that recurrent seizures might lead to greater neuronal injury in the younger population, or the effect might be harder to detect in the older people who have higher background plasma levels of these proteins.

Among patients with epilepsy, we did not find any significant differences in clinical variables, in patients with high plasma NfL levels (>95^th^ percentile) versus those with lower levels, and by NfL and GFAP quartiles (Supplementary Material). Of note, GFAP levels, NfL levels and age-adjusted NfL z-scores, were not different between lesional epilepsy and non-lesional epilepsy, or between different lesion types, such as gliosis or stroke. In a recent publication that found a small proportion of patients with very high biomarker levels for NfL, the majority of patients with elevated NfL had had a previous stroke(31), and there is strong evidence for persistently elevated plasma NfL levels in post-stroke epilepsy(22). Our findings are therefore important in this context, since we found elevated levels in patients who did not have post-stroke epilepsy. There is a known relationship between increasing age and increased NfL levels(28). However, in our study, patients with very elevated plasma NfL levels, were in fact younger than those with a lower NfL level. This further suggests that elevated levels are more likely to be related to epilepsy than non-epilepsy and age-related factors such as white matter disease or comorbid neurodegenerative disease.

There has been limited research examining the relationship between seizure frequency and blood biomarkers (31). We did not identify a statistically significant relationship between seizure frequency and plasma NfL levels. In our study, a high seizure burden was noted in both patients with highly elevated and more modest plasma NfL levels (average 1-3 seizures per month in both cohorts). This may suggest that seizures frequency is not a clear indicator of neuronal damage, or plasma NfL may lack sensitivity in detecting neuronal damage associated with seizure frequency. However, it is also recognised that seizure frequency may not be reliably reported by patients(32). Further research using more reliable seizure detection methods (e.g., wearable or subscalp/intracranial devices) and correlation with the timing of seizures is needed.

The strengths of our study include its relatively large sample size among studies examining the utility of biomarkers in epilepsy, with diagnosis of each patient confirmed after a multidisciplinary discussion of comprehensive inpatient assessments including video-EEG monitoring and formal neuropsychiatric assessment. The study is limited by the relatively small size of the subgroups which impacts on the ability to identify differences between subgroups of patients with epilepsy. The relationship between age and GFAP is poorly understood, however there is some evidence to that GFAP also increases with age(33,34). As a reference cohort for GFAP levels is not available, we were unable to perform GAMLSS modelling for GFAP. Future research should focus on developing age stratified reference ranges for GFAP. Where possible, data was cross-referenced with the patient’s medical record, but there are inherent limitations of such a retrospective study. Covariates known to influence biomarkers such as weight and renal function, and variables that might affect their levels (such as antiseizure medications, sample time after/before most recent seizure), should also be explored in future studies that include larger subgroups. Finally, future research should investigate rate of change in the levels of biomarkers over time, their associations with seizures in the short-medium term (e.g., days to months), and over the longer term (e.g., years).

In conclusion, our findings suggest that finding an elevated plasma NfL or GFAP level in an individual patient may imply an underlying epilepsy diagnosis, and caution against a diagnosis of PNES alone. This is particularly the case in younger individuals with highly elevated levels. Further examination in larger sample sizes of the association between NfL and GFAP levels and specific epilepsy subtypes or seizure characteristics, may provide valuable insights into disease heterogeneity and contribute to the refinement of diagnosis, understanding pathophysiological mechanisms, and formulating treatment approaches.

## Data Availability

Data produced in the present study are available upon reasonable request to the authors

## Acknowledgements

This study was supported by MACH MRFF RART 2.2, NHMRC (1185180). The role of these funding sources was to support research study staff and biosample analyses.

**Hannah Dobson:** Conceptualisation, Data curation, Writing – original draft, **Said Al Maawali:** Conceptualisation, Data curation, Writing – original draft, **Charles Malpas:** Formal analysis, Writing – review & editing**; Alexander F Santillo:** Writing – Review & Editing**; Matthew Kang:** Data curation, Writing – Review & Editing, **Marian Todaro:** Data curation, Writing – Review & Editing, **Rosie Watson:** Investigation, Writing – Review & Editing, **Nawaf Yassi:** Investigation, Writing – Review & Editing, **Kaj Blennow:** Writing – Review & Editing, **Henrik Zetterberg:** Writing – Review & Editing, **Emma Foster:** Writing – Review & Editing, **Andrew Neal:** Writing – Review & Editing, **Dennis Velakoulis:** Conceptualisation, Methodology, Writing –Review & Editing, Funding acquisition, **Terence John O’Brien:** Conceptualisation, Methodology, Writing –Review & Editing, Supervision, Funding acquisition, **Dhamidhu Eratne:** Conceptualisation, Methodology, Formal analysis, Data curation, Writing – Original draft, Supervision, Funding acquisition**; Patrick Kwan:** Conceptualisation, Methodology, Writing –Review & Editing, Supervision, Funding acquisition

Finally, the authors thank all the participants and their families for their participation, and the other clinicians who contribute to The MiND Study Group.

## Appendix 1: Coinvestigators

On behalf of others in The MiND Study Group:

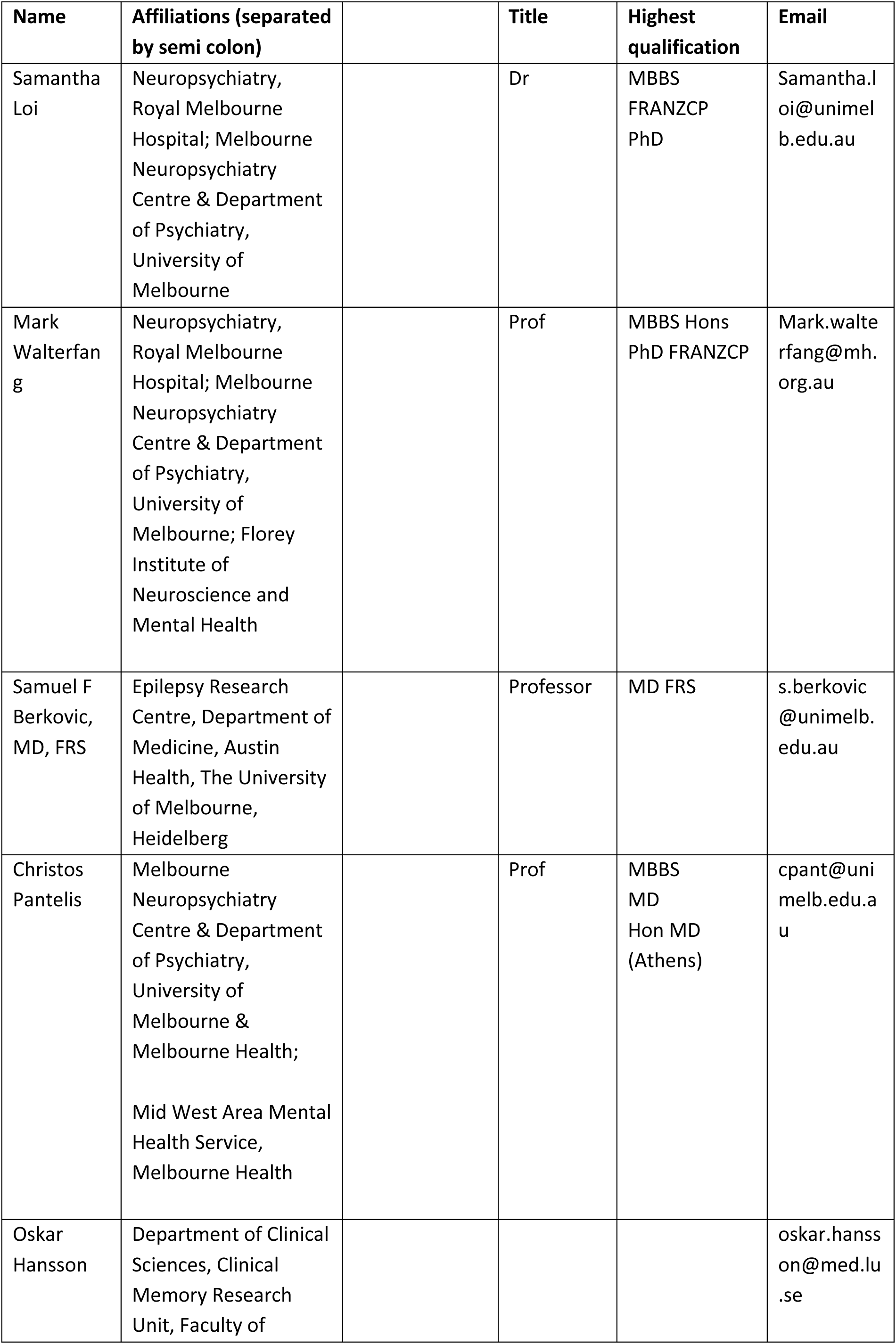
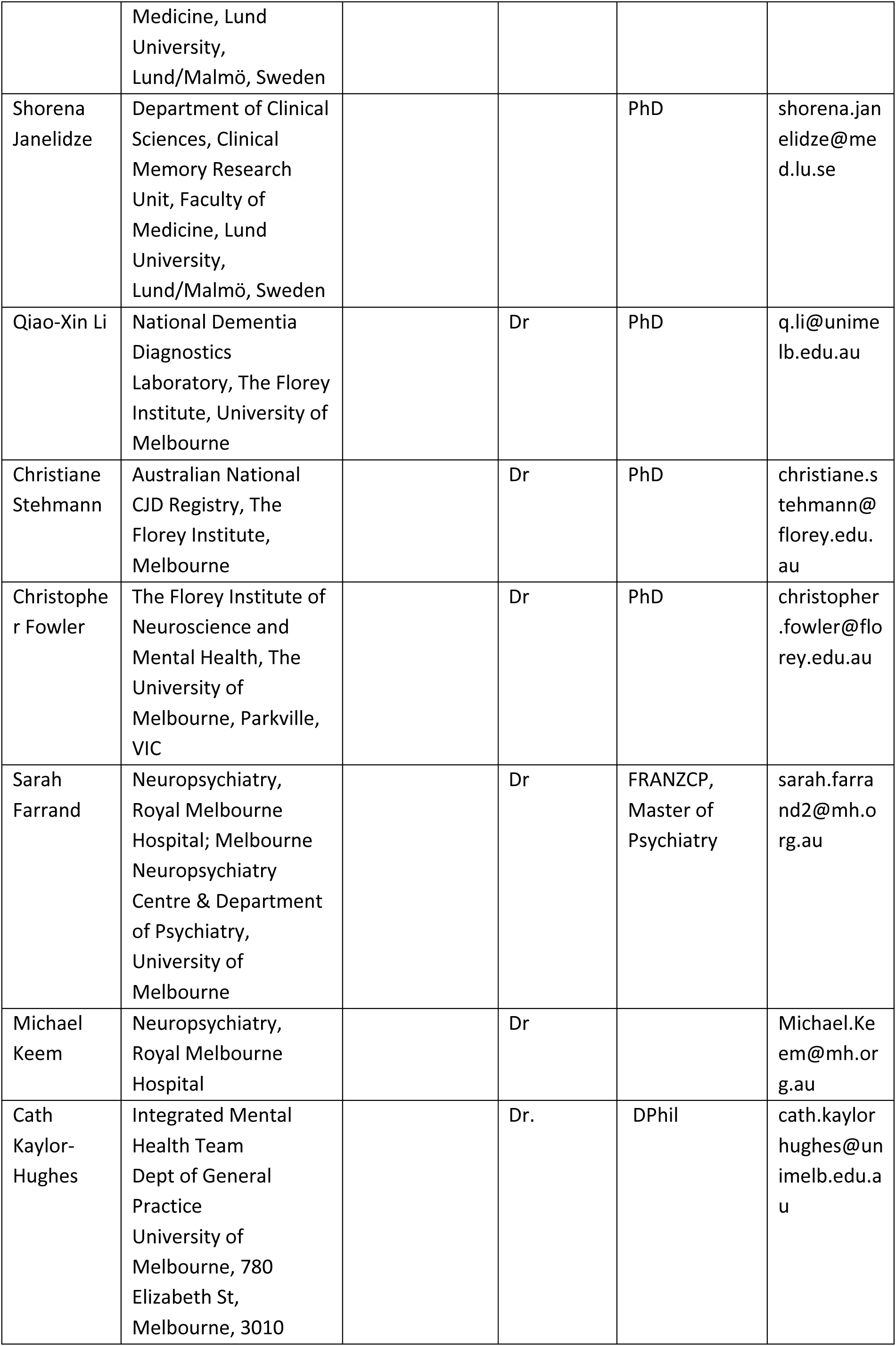
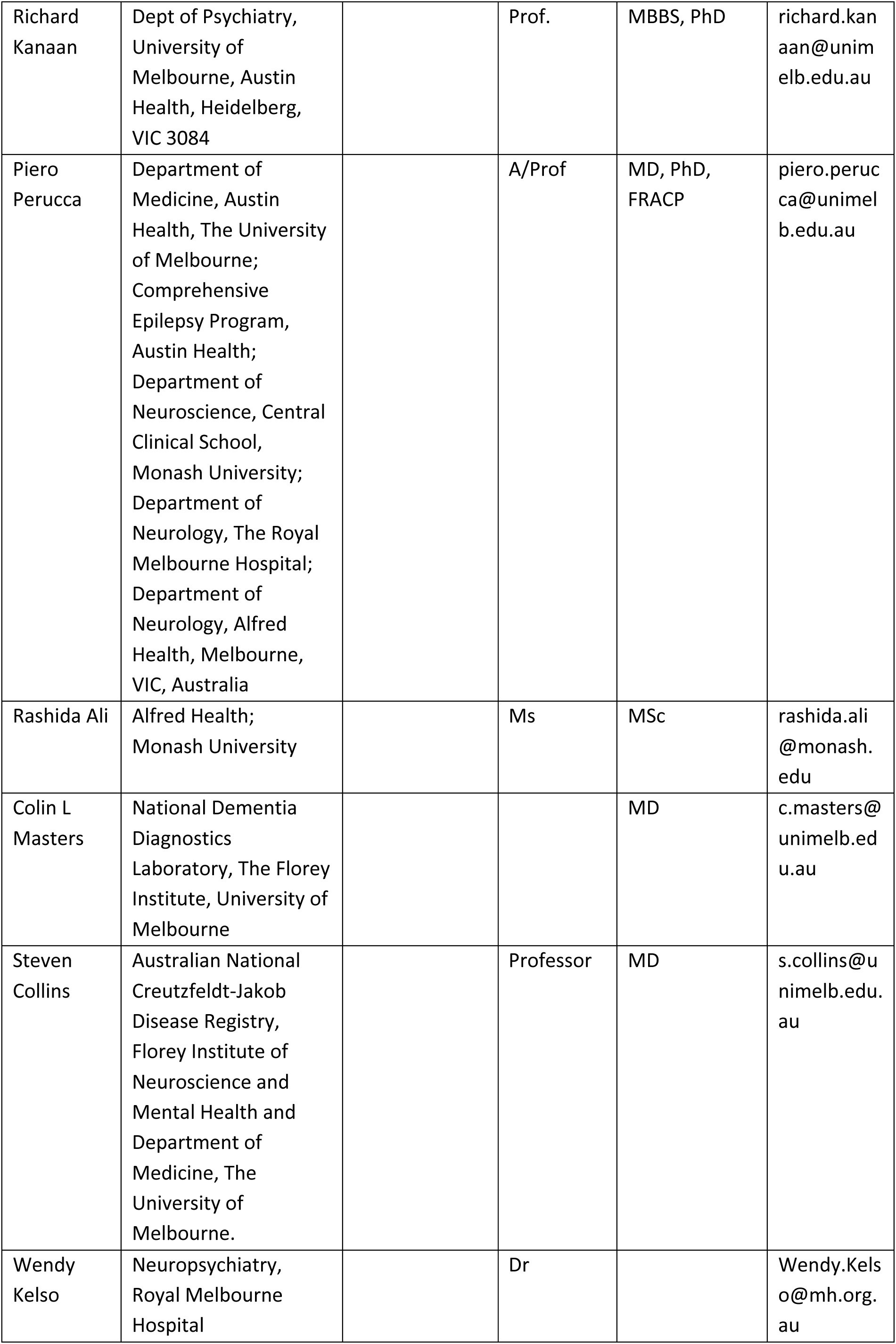
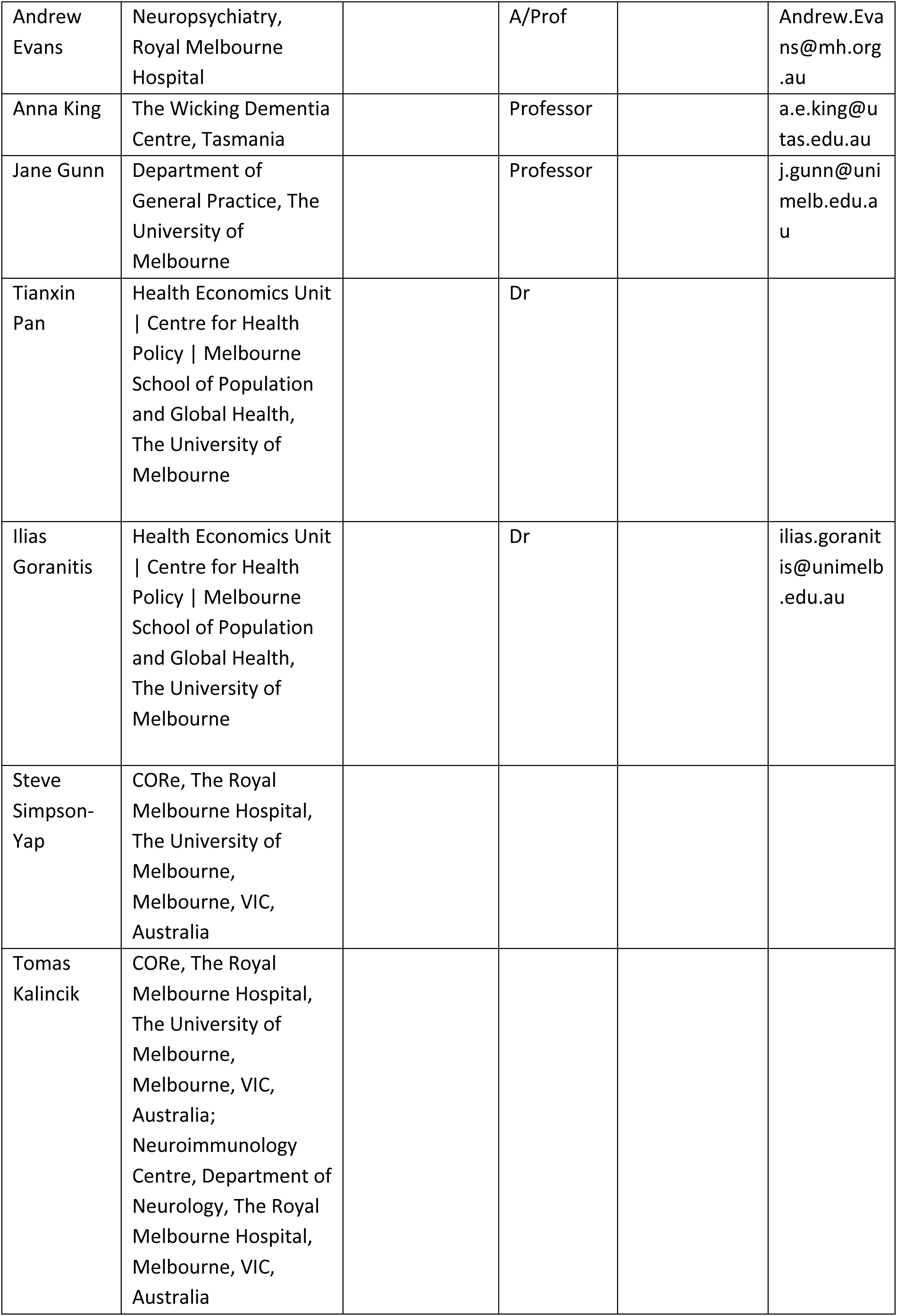

## LIST OF REFERENCES

1. Fisher RS, Acevedo C, Arzimanoglou A, Bogacz A, Cross JH, Elger CE, et al. ILAE official report: a practical clinical definition of epilepsy. Epilepsia. 2014;55(4):475–82.

2. Krumholz A, Wiebe S, Gronseth GS, Gloss DS, Sanchez AM, Kabir AA, et al. Evidence-based guideline: management of an unprovoked first seizure in adults: report of the guideline development subcommittee of the American Academy of Neurology and the American Epilepsy Society: evidence-based guideline. Epilepsy currents. 2015;15(3):144–52.

3. Foster E, Holper S, Chen Z, Kwan P. Presentation and management of community-onset vs hospital-onset first seizures. Neurol Clin Pract. 2018 Oct;8(5):421–8.

4. Holper S, Foster E, Chen Z, Kwan P. Emergency presentation of new onset versus recurrent undiagnosed seizures: A retrospective review. Emergency Medicine Australasia. 2020;32(3):430–7.

5. Kanemoto K, LaFrance WC, Duncan R, Gigineishvili D, Park SP, Tadokoro Y, et al. PNES around the world: Where we are now and how we can close the diagnosis and treatment gaps-an ILAE PNES Task Force report. Epilepsia Open. 2017 Sep;2(3):307–16.

6. Ghougassian DF, d’Souza W, Cook MJ, O’Brien TJ. Evaluating the utility of inpatient video-EEG monitoring. Epilepsia. 2004 Aug;45(8):928–32.

7. Bodde NMG, Brooks JL, Baker GA, Boon PAJM, Hendriksen JGM, Mulder OG, et al. Psychogenic non-epileptic seizures—Definition, etiology, treatment and prognostic issues: A critical review. Seizure. 2009 Oct 1;18(8):543–53.

8. Sirven JI, Glosser DS. Psychogenic nonepileptic seizures: theoretic and clinical considerations. Neuropsychiatry Neuropsychol Behav Neurol. 1998 Oct;11(4):225–35.

9. Tao G, Auvrez C, Nightscales R, Barnard S, McCartney L, Malpas CB, et al. Association Between Psychiatric Comorbidities and Mortality in Epilepsy. Neurol Clin Pract. 2021 Oct;11(5):429–37.

10. Foster E, Chen Z, Zomer E, Rychkova M, Carney P, O’Brien TJ, et al. The costs of epilepsy in Australia: A productivity-based analysis. Neurology. 2020 Dec 15;95(24):e3221–31.

11. Gaetani L, Blennow K, Calabresi P, Di Filippo M, Parnetti L, Zetterberg H. Neurofilament light chain as a biomarker in neurological disorders. Journal of Neurology, Neurosurgery & Psychiatry. 2019 Aug;90(8):870–81.

12. Abdelhak A, Foschi M, Abu-Rumeileh S, Yue JK, D’Anna L, Huss A, et al. Blood GFAP as an emerging biomarker in brain and spinal cord disorders. Nat Rev Neurol. 2022 Mar;18(3):158–72.

13. Eratne D, Loi SM, Walia N, Farrand S, Li QX, Varghese S, et al. A pilot study of the utility of cerebrospinal fluid neurofilament light chain in differentiating neurodegenerative from psychiatric disorders: A ‘C-reactive protein’ for psychiatrists and neurologists? Australian & New Zealand Journal of Psychiatry. 2020;54(1):57–67.

14. Eratne D, Keem M, Lewis C, Kang M, Walterfang M, Farrand S, et al. Cerebrospinal fluid neurofilament light chain differentiates behavioural variant frontotemporal dementia progressors from non-progressors. Journal of the Neurological Sciences. 2022;442:120439.

15. Kang MJ, Eratne D, Dobson H, Malpas CB, Keem M, Lewis C, et al. Cerebrospinal fluid neurofilament light predicts longitudinal diagnostic change in patients with psychiatric and neurodegenerative disorders. Acta Neuropsychiatrica. 2023;1–12.

16. Eratne D, Kang M, Malpas C, Simpson-Yap S, Lewis C, Dang C, et al. Plasma neurofilament light in behavioural variant frontotemporal dementia compared to mood and psychotic disorders. Aust N Z J Psychiatry. 2023 Jul 21;48674231187312.

17. Caciagli L, Bernasconi A, Wiebe S, Koepp MJ, Bernasconi N, Bernhardt BC. A meta-analysis on progressive atrophy in intractable temporal lobe epilepsy: Time is brain? Neurology. 2017 Aug 1;89(5):506–16.

18. Galovic M, de Tisi J, McEvoy AW, Miserocchi A, Vos SB, Borzi G, et al. Resective surgery prevents progressive cortical thinning in temporal lobe epilepsy. Brain. 2020 Dec 5;143(11):3262–72.

19. Banote RK, Akel S, Zelano J. Blood biomarkers in epilepsy. Acta Neurologica Scandinavica. 2022;146(4):362–8.

20. Giovannini G, Bedin R, Ferraro D, Vaudano AE, Mandrioli J, Meletti S. Serum neurofilament light as biomarker of seizure-related neuronal injury in status epilepticus. Epilepsia [Internet]. 2022 Jan [cited 2022 Mar 30];63(1). Available from: https://onlinelibrary.wiley.com/doi/10.1111/epi.17132

21. Giovannini G, Bedin R, Orlandi N, Turchi G, Cioclu MC, Biagioli N, et al. Neuro-glial degeneration in Status Epilepticus: Exploring the role of serum levels of Neurofilament light chains and S100B as prognostic biomarkers for short-term functional outcome. Epilepsy & Behavior. 2023;140:109131.

22. Dev P, Cyriac M, Chakravarty K, Pathak A. Blood and CSF biomarkers for post-stroke epilepsy: a systematic review. Acta Epileptologica. 2022 Jul 1;4(1):21.

23. Nass RD, Akgün K, Dague KO, Elger CE, Reichmann H, Ziemssen T, et al. CSF and Serum Biomarkers of Cerebral Damage in Autoimmune Epilepsy. Front Neurol. 2021;12:647428.

24. Ueda M, Suzuki M, Hatanaka M, Nakamura T, Hirayama M, Katsuno M. Serum neurofilament light chain in patients with epilepsy and cognitive impairment. Epileptic Disorders. 2023;

25. Mochol M, Taubøll E, Aukrust P, Ueland T, Andreassen OA, Svalheim S. Serum Markers of Neuronal Damage and Astrocyte Activity in Patients with Chronic Epilepsy: Elevated Levels of Glial Fibrillary Acidic Protein. Acta Neurologica Scandinavica. 2023;2023.

26. Elhady M, Youness ER, AbuShady MM, Nassar MS, Elaziz AA, Masoud MM, et al. Circulating glial fibrillary acidic protein and ubiquitin carboxy-terminal hydrolase-L1 as markers of neuronal damage in children with epileptic seizures. Child’s Nervous System. 2021;37:879–84.

27. Simani L, Elmi M, Asadollahi M. Serum GFAP level: a novel adjunctive diagnostic test in differentiate epileptic seizures from psychogenic attacks. Seizure. 2018;61:41–4.

28. Simrén J, Andreasson U, Gobom J, Suarez Calvet M, Borroni B, Gillberg C, et al. Establishment of reference values for plasma neurofilament light based on healthy individuals aged 5–90 years. Brain Commun. 2022 Jul 4;4(4):fcac174.

29. So EL, Radhakrishnan K, Silbert PL, Cascino GD, Sharbrough FW, O’Brien PC. Assessing changes over time in temporal lobectomy: outcome by scoring seizure frequency. Epilepsy Res. 1997 May;27(2):119– 25.

30. Alfred Health. Neuroscience Bio-databank [Internet]. [cited 2023 Aug 21]. Available from: https://www.alfredhealth.org.au/research/research-areas/ANB-Landing

31. Akel S, Asztely F, Banote RK, Axelsson M, Zetterberg H, Zelano J. Neurofilament light, glial fibrillary acidic protein, and tau in a regional epilepsy cohort: High plasma levels are rare but related to seizures. Epilepsia. 2023 Jul 19;

32. Rayner G, Jackson GD, Wilson SJ. Mechanisms of memory impairment in epilepsy depend on age at disease onset. Neurology. 2016 Oct 18;87(16):1642–9.

33. Vågberg M, Norgren N, Dring A, Lindqvist T, Birgander R, Zetterberg H, et al. Levels and Age Dependency of Neurofilament Light and Glial Fibrillary Acidic Protein in Healthy Individuals and Their Relation to the Brain Parenchymal Fraction. PLoS One. 2015;10(8):e0135886.

34. Kohama SG, Goss JR, Finch CE, McNeill TH. Increases of glial fibrillary acidic protein in the aging female mouse brain. Neurobiol Aging. 1995;16(1):59–67.

35. Dittrich A, Ashton NJ, Zetterberg H, Blennow K, Zettergren A, Simrén J, et al. Association of Chronic Kidney Disease With Plasma NfL and Other Biomarkers of Neurodegeneration: The H70 Birth Cohort Study in Gothenburg. Neurology. 2023 May 24;10.1212/WNL.0000000000207419.

36. Polymeris AA, Helfenstein F, Benkert P, Aeschbacher S, Leppert D, Coslovsky M, et al. Renal Function and Body Mass Index Contribute to Serum Neurofilament Light Chain Levels in Elderly Patients With Atrial Fibrillation. Front Neurosci. 2022 Apr 14;16:819010.

37. Eratne D, Janelidze S, Malpas CB, Loi S, Walterfang M, Merritt A, et al. Plasma neurofilament light chain protein is not increased in treatment-resistant schizophrenia and first-degree relatives. Australian & New Zealand Journal of Psychiatry. 2022;56(10):1295–305.

