## Supplementary Material for "Elevated plasma neurofilament light & glial fibrillary acidic protein in epilepsy versus non-epileptic seizures & non-epileptic disorders"

### Supplementary Table 1: Summary descriptives table by groups of `nfl.quartile'

| | [ALL] | Q1 | Q2 | Q3 | Q4 | p.overall | N |
|---|---|---|---|---|---|---|---|
| | *N=115* | *N=29* | *N=29* | *N=28* | *N=29* | | |
| age | 40.8 (15.1) | 32.1 (8.28) | 36.9 (10.3) | 44.7 (12.8) | 49.8 (19.9) | <0.001 | 115 |
| sex: | | | | | | 0.605 | 115 |
| Female | 63 (54.8%) | 16 (55.2%) | 18 (62.1%) | 16 (57.1%) | 13 (44.8%) | | |
| Male | 52 (45.2%) | 13 (44.8%) | 11 (37.9%) | 12 (42.9%) | 16 (55.2%) | | |
| nfl | 12.4 (16.5) | 3.94 (0.70) | 6.09 (0.79) | 9.37 (1.40) | 30.2 (25.4) | <0.001 | 115 |
| lognfl | 0.93 (0.33) | 0.59 (0.08) | 0.78 (0.06) | 0.97 (0.06) | 1.38 (0.27) | <0.001 | 115 |
| nfl.z | 0.76 (1.36) | -0.47 (0.82) | 0.44 (0.83) | 0.97 (0.86) | 2.09 (1.38) | <0.001 | 115 |
| percentile | 0.66 (0.31) | 0.36 (0.26) | 0.63 (0.25) | 0.77 (0.23) | 0.88 (0.21) | <0.001 | 115 |
| gfap | 107 (116) | 63.5 (54.9) | 57.1 (21.4) | 151 (192) | 158 (77.6) | <0.001 | 115 |
| loggfap | 1.90 (0.30) | 1.72 (0.23) | 1.72 (0.17) | 2.00 (0.34) | 2.15 (0.20) | <0.001 | 115 |
| Seizure Localisation: | | | | | | 0.632 | 115 |
| FLE | 15 (13.0%) | 6 (20.7%) | 1 (3.45%) | 5 (17.9%) | 3 (10.3%) | | |
| IGE | 9 (7.83%) | 2 (6.90%) | 3 (10.3%) | 2 (7.14%) | 2 (6.90%) | | |

| | [ALL] | Q1 | Q2 | Q3 | Q4 | p.overall | N |
|---|---|---|---|---|---|---|---|
| | *N=115* | *N=29* | *N=29* | *N=28* | *N=29* | | |
| TLE | 49 (42.6%) | 13 (44.8%) | 15 (51.7%) | 11 (39.3%) | 10 (34.5%) | | |
| Unclassified | 37 (32.2%) | 7 (24.1%) | 10 (34.5%) | 9 (32.1%) | 11 (37.9%) | | |
| Unknown | 5 (4.35%) | 1 (3.45%) | 0 (0.00%) | 1 (3.57%) | 3 (10.3%) | | |
| Age of Seizure Onset | 19.9 (17.4) | 18.7 (11.6) | 17.8 (13.0) | 16.4 (17.1) | 26.7 (24.0) | 0.102 | 114 |
| Seizure Duration (Years) | 21.1 (17.5) | 13.5 (11.2) | 19.6 (13.0) | 28.4 (19.1) | 23.0 (21.9) | 0.011 | 115 |
| Seizure Frequency | 7.22 (1.97) | 7.21 (1.69) | 7.24 (1.98) | 7.09 (2.54) | 7.35 (1.67) | 0.977 | 99 |
| Seizure Type | | | | | | 0.580 | 102 |
| Focal | 69 (67.6%) | 18 (72.0%) | 16 (55.2%) | 18 (75.0%) | 17 (70.8%) | | |
| FTBTC | 17 (16.7%) | 4 (16.0%) | 6 (20.7%) | 2 (8.33%) | 5 (20.8%) | | |
| Generalised | 16 (15.7%) | 3 (12.0%) | 7 (24.1%) | 4 (16.7%) | 2 (8.33%) | | |
| Seizure Type, further categories | | | | | | 0.644 | 101 |
| FAS | 17 (16.8%) | 4 (16.0%) | 3 (10.7%) | 5 (20.8%) | 5 (20.8%) | | |
| FIAS | 44 (43.6%) | 10 (40.0%) | 12 (42.9%) | 10 (41.7%) | 12 (50.0%) | | |
| Focal | 8 (7.92%) | 4 (16.0%) | 1 (3.57%) | 3 (12.5%) | 0 (0.00%) | | |
| FTBTC | 17 (16.8%) | 4 (16.0%) | 6 (21.4%) | 2 (8.33%) | 5 (20.8%) | | |

| | [ALL] | Q1 | Q2 | Q3 | Q4 | p.overall | N |
|---|---|---|---|---|---|---|---|
| | *N=115* | *N=29* | *N=29* | *N=28* | *N=29* | | |
| Generalised | 15 (14.9%) | 3 (12.0%) | 6 (21.4%) | 4 (16.7%) | 2 (8.33%) | | |
| MRI Abnormality | | | | | | 0.482 | 89 |
| Abnormal | 60 (67.4%) | 18 (72.0%) | 16 (72.7%) | 10 (52.6%) | 16 (69.6%) | | |
| Normal | 29 (32.6%) | 7 (28.0%) | 6 (27.3%) | 9 (47.4%) | 7 (30.4%) | | |
| MRI Lesion | | | | | | 0.321 | 89 |
| No | 53 (59.6%) | 12 (48.0%) | 12 (54.5%) | 14 (73.7%) | 15 (65.2%) | | |
| Yes | 36 (40.4%) | 13 (52.0%) | 10 (45.5%) | 5 (26.3%) | 8 (34.8%) | | |
| MRI Lesion Type | | | | | | 0.491 | 36 |
| Cavernoma/vascular malformation | 4 (11.1%) | 2 (15.4%) | 0 (0.00%) | 1 (20.0%) | 1 (12.5%) | | |
| Gliosis/stroke | 2 (5.56%) | 0 (0.00%) | 1 (10.0%) | 1 (20.0%) | 0 (0.00%) | | |
| Hippocampal sclerosis | 8 (22.2%) | 2 (15.4%) | 3 (30.0%) | 1 (20.0%) | 2 (25.0%) | | |
| Hippocampal sclerosis + vascular | 2 (5.56%) | 1 (7.69%) | 0 (0.00%) | 0 (0.00%) | 1 (12.5%) | | |
| MCD/FCD | 9 (25.0%) | 2 (15.4%) | 2 (20.0%) | 2 (40.0%) | 3 (37.5%) | | |
| Other | 2 (5.56%) | 0 (0.00%) | 2 (20.0%) | 0 (0.00%) | 0 (0.00%) | | |
| Tumour/glioma | 9 (25.0%) | 6 (46.2%) | 2 (20.0%) | 0 (0.00%) | 1 (12.5%) | | |

GFAP: glial fibrillary acidic protein; NfL: neurofilament light chain protein; FLE: frontal lobe epilepsy; IGE: idiopathic generalised epilepsy; TLE: temporal lobe epilepsy; FTBTC: focal to bilateral tonic clonic; MCD: malformation of cortical development; FCD: focal cortical dysplasia

**Supplementary Table 2: Summary descriptives table by groups of `gfap.quartile'**

| | [ALL] | Q1 | Q2 | Q3 | Q4 | p.overall | N |
|---|---|---|---|---|---|---|---|
| | *N=115* | *N=29* | *N=29* | *N=28* | *N=29* | | |
| age | 40.8 (15.1) | 33.2 (10.1) | 38.9 (11.8) | 46.3 (13.6) | 45.2 (19.8) | 0.002 | 115 |
| sex: | | | | | | 0.111 | 115 |
| Female | 63 (54.8%) | 13 (44.8%) | 21 (72.4%) | 16 (57.1%) | 13 (44.8%) | | |
| Male | 52 (45.2%) | 16 (55.2%) | 8 (27.6%) | 12 (42.9%) | 16 (55.2%) | | |
| nfl | 12.4 (16.5) | 5.03 (1.45) | 6.62 (2.84) | 12.1 (9.05) | 26.0 (27.0) | <0.001 | 115 |
| lognfl | 0.93 (0.33) | 0.68 (0.13) | 0.78 (0.19) | 1.01 (0.25) | 1.25 (0.37) | <0.001 | 115 |
| nfl.z | 0.76 (1.36) | 0.10 (0.84) | 0.23 (1.15) | 0.97 (1.15) | 1.74 (1.56) | <0.001 | 115 |
| percentile | 0.66 (0.31) | 0.53 (0.27) | 0.55 (0.32) | 0.74 (0.27) | 0.81 (0.28) | <0.001 | 115 |
| gfap | 107 (116) | 38.0 (8.12) | 60.4 (5.96) | 88.2 (12.6) | 241 (167) | <0.001 | 115 |
| loggfap | 1.90 (0.30) | 1.57 (0.10) | 1.78 (0.04) | 1.94 (0.06) | 2.31 (0.23) | <0.001 | 115 |
| Seizure Localisation: | | | | | | 0.180 | 115 |
| FLE | 15 (13.0%) | 5 (17.2%) | 2 (6.90%) | 1 (3.57%) | 7 (24.1%) | | |
| IGE | 9 (7.83%) | 5 (17.2%) | 1 (3.45%) | 1 (3.57%) | 2 (6.90%) | | |
| TLE | 49 (42.6%) | 12 (41.4%) | 14 (48.3%) | 15 (53.6%) | 8 (27.6%) | | |

| | [ALL] *N=115* | Q1 *N=29* | Q2 *N=29* | Q3 *N=28* | Q4 *N=29* | p.overall | N |
|---|---|---|---|---|---|---|---|
| Unclassified | 37 (32.2%) | 7 (24.1%) | 10 (34.5%) | 9 (32.1%) | 11 (37.9%) | | |
| Unknown | 5 (4.35%) | 0 (0.00%) | 2 (6.90%) | 2 (7.14%) | 1 (3.45%) | | |
| Age Seizure Onset | 19.9 (17.4) | 19.0 (13.1) | 17.5 (11.1) | 21.5 (22.3) | 21.8 (20.9) | 0.755 | 114 |
| Seizure Duration (years) | 21.1 (17.5) | 14.8 (12.0) | 21.4 (16.7) | 24.8 (20.9) | 23.4 (18.5) | 0.136 | 115 |
| Seizure Frequency | 7.22 (1.97) | 7.12 (2.41) | 7.17 (1.52) | 7.54 (2.11) | 7.08 (1.78) | 0.840 | 99 |
| Seizure Type | | | | | | 0.492 | 102 |
| Focal | 69 (67.6%) | 15 (57.7%) | 18 (72.0%) | 18 (75.0%) | 18 (66.7%) | | |
| FTBTC | 17 (16.7%) | 4 (15.4%) | 5 (20.0%) | 2 (8.33%) | 6 (22.2%) | | |
| Generalised | 16 (15.7%) | 7 (26.9%) | 2 (8.00%) | 4 (16.7%) | 3 (11.1%) | | |
| Seizure Type, further categories | | | | | | 0.776 | 101 |
| FAS | 17 (16.8%) | 5 (20.0%) | 2 (8.00%) | 5 (20.8%) | 5 (18.5%) | | |
| FIAS | 44 (43.6%) | 8 (32.0%) | 13 (52.0%) | 12 (50.0%) | 11 (40.7%) | | |
| Focal | 8 (7.92%) | 2 (8.00%) | 3 (12.0%) | 1 (4.17%) | 2 (7.41%) | | |
| FTBTC | 17 (16.8%) | 4 (16.0%) | 5 (20.0%) | 2 (8.33%) | 6 (22.2%) | | |
| Generalised | 15 (14.9%) | 6 (24.0%) | 2 (8.00%) | 4 (16.7%) | 3 (11.1%) | | |

| | [ALL] *N=115* | Q1 *N=29* | Q2 *N=29* | Q3 *N=28* | Q4 *N=29* | p.overall | N |
|---|---|---|---|---|---|---|---|
| MRI Abnormality | | | | | | 0.624 | 89 |
| Abnormal | 60 (67.4%) | 16 (72.7%) | 15 (65.2%) | 15 (75.0%) | 14 (58.3%) | | |
| Normal | 29 (32.6%) | 6 (27.3%) | 8 (34.8%) | 5 (25.0%) | 10 (41.7%) | | |
| MRI Lesion | | | | | | 0.718 | 89 |
| No | 53 (59.6%) | 14 (63.6%) | 12 (52.2%) | 11 (55.0%) | 16 (66.7%) | | |
| Yes | 36 (40.4%) | 8 (36.4%) | 11 (47.8%) | 9 (45.0%) | 8 (33.3%) | | |
| MRI Lesion Type | | | | | | 0.129 | 36 |
| Cavernoma/vascular malformation | 4 (11.1%) | 1 (12.5%) | 1 (9.09%) | 2 (22.2%) | 0 (0.00%) | | |
| Gliosis/stroke | 2 (5.56%) | 1 (12.5%) | 0 (0.00%) | 1 (11.1%) | 0 (0.00%) | | |
| Hippocampal sclerosis | 8 (22.2%) | 0 (0.00%) | 4 (36.4%) | 3 (33.3%) | 1 (12.5%) | | |
| Hippocampal sclerosis + vascular | 2 (5.56%) | 0 (0.00%) | 1 (9.09%) | 0 (0.00%) | 1 (12.5%) | | |
| MCD/FCD | 9 (25.0%) | 1 (12.5%) | 3 (27.3%) | 0 (0.00%) | 5 (62.5%) | | |
| Other | 2 (5.56%) | 1 (12.5%) | 0 (0.00%) | 1 (11.1%) | 0 (0.00%) | | |
| Tumour/glioma | 9 (25.0%) | 4 (50.0%) | 2 (18.2%) | 2 (22.2%) | 1 (12.5%) | | |

GFAP: glial fibrillary acidic protein; NfL: neurofilament light chain protein; FLE: frontal lobe epilepsy; IGE: idiopathic generalised epilepsy; TLE: temporal lobe epilepsy; FTBTC: focal to bilateral tonic clonic; MCD: malformation of cortical development; FCD: focal cortical dysplasia

**Supplementary Table 3: Summary descriptives table by groups of `nfl.z.quartile'**

| | [ALL] | Q1 | Q2 | Q3 | Q4 | p.overall | N |
|---|---|---|---|---|---|---|---|
| | *N=115* | *N=29* | *N=29* | *N=28* | *N=29* | | |
| age | 40.8 (15.1) | 46.7 (14.7) | 42.7 (17.1) | 39.4 (15.4) | 34.6 (10.3) | 0.017 | 115 |
| sex: | | | | | | 0.493 | 115 |
| Female | 63 (54.8%) | 18 (62.1%) | 17 (58.6%) | 12 (42.9%) | 16 (55.2%) | | |
| Male | 52 (45.2%) | 11 (37.9%) | 12 (41.4%) | 16 (57.1%) | 13 (44.8%) | | |
| nfl | 12.4 (16.5) | 6.07 (4.24) | 7.68 (6.29) | 9.09 (4.78) | 26.8 (27.1) | <0.001 | 115 |
| lognfl | 0.93 (0.33) | 0.71 (0.23) | 0.81 (0.22) | 0.91 (0.20) | 1.28 (0.35) | <0.001 | 115 |
| nfl.z | 0.76 (1.36) | -0.84 (0.45) | 0.20 (0.30) | 1.11 (0.31) | 2.57 (0.79) | <0.001 | 115 |
| percentile | 0.66 (0.31) | 0.22 (0.12) | 0.58 (0.11) | 0.86 (0.06) | 0.98 (0.01) | <0.001 | 115 |
| gfap | 107 (116) | 81.7 (60.7) | 69.4 (64.6) | 93.5 (68.9) | 183 (183) | <0.001 | 115 |
| loggfap | 1.90 (0.30) | 1.83 (0.25) | 1.75 (0.26) | 1.89 (0.26) | 2.13 (0.32) | <0.001 | 115 |
| localisation: | | | | | | 0.319 | 115 |
| FLE | 15 (13.0%) | 2 (6.90%) | 6 (20.7%) | 2 (7.14%) | 5 (17.2%) | | |
| IGE | 9 (7.83%) | 2 (6.90%) | 4 (13.8%) | 1 (3.57%) | 2 (6.90%) | | |
| TLE | 49 (42.6%) | 16 (55.2%) | 11 (37.9%) | 14 (50.0%) | 8 (27.6%) | | |

| | [ALL] | Q1 | Q2 | Q3 | Q4 | p.overall | N |
|---|---|---|---|---|---|---|---|
| | *N=115* | *N=29* | *N=29* | *N=28* | *N=29* | | |
| Unclassified | 37 (32.2%) | 8 (27.6%) | 8 (27.6%) | 8 (28.6%) | 13 (44.8%) | | |
| Unknown | 5 (4.35%) | 1 (3.45%) | 0 (0.00%) | 3 (10.7%) | 1 (3.45%) | | |
| age_onset | 19.9 (17.4) | 26.5 (21.2) | 20.2 (17.8) | 17.5 (16.3) | 15.5 (11.7) | 0.086 | 114 |
| sz_duration | 21.1 (17.5) | 20.2 (17.6) | 22.4 (18.1) | 22.5 (20.2) | 19.1 (14.4) | 0.853 | 115 |
| sz_frequency | 7.22 (1.97) | 7.04 (2.14) | 7.32 (2.32) | 7.17 (1.40) | 7.38 (2.00) | 0.924 | 99 |
| sz_type: | | | | | | 0.445 | 102 |
| Focal | 69 (67.6%) | 21 (75.0%) | 11 (47.8%) | 18 (75.0%) | 19 (70.4%) | | |
| FTBTC | 17 (16.7%) | 3 (10.7%) | 6 (26.1%) | 3 (12.5%) | 5 (18.5%) | | |
| Generalised | 16 (15.7%) | 4 (14.3%) | 6 (26.1%) | 3 (12.5%) | 3 (11.1%) | | |
| sz_type_2: | | | | | | 0.775 | 101 |
| FAS | 17 (16.8%) | 5 (17.9%) | 1 (4.55%) | 5 (20.8%) | 6 (22.2%) | | |
| FIAS | 44 (43.6%) | 13 (46.4%) | 9 (40.9%) | 10 (41.7%) | 12 (44.4%) | | |
| Focal | 8 (7.92%) | 3 (10.7%) | 1 (4.55%) | 3 (12.5%) | 1 (3.70%) | | |
| FTBTC | 17 (16.8%) | 3 (10.7%) | 6 (27.3%) | 3 (12.5%) | 5 (18.5%) | | |
| Generalised | 15 (14.9%) | 4 (14.3%) | 5 (22.7%) | 3 (12.5%) | 3 (11.1%) | | |

| | [ALL] | Q1 | Q2 | Q3 | Q4 | p.overall | N |
|---|---|---|---|---|---|---|---|
| | *N=115* | *N=29* | *N=29* | *N=28* | *N=29* | | |
| mri_abnormal: | | | | | | 0.097 | 89 |
| Abnormal | 60 (67.4%) | 17 (73.9%) | 18 (81.8%) | 10 (47.6%) | 15 (65.2%) | | |
| Normal | 29 (32.6%) | 6 (26.1%) | 4 (18.2%) | 11 (52.4%) | 8 (34.8%) | | |
| mri_lesional: | | | | | | 0.327 | 89 |
| No | 53 (59.6%) | 12 (52.2%) | 13 (59.1%) | 16 (76.2%) | 12 (52.2%) | | |
| Yes | 36 (40.4%) | 11 (47.8%) | 9 (40.9%) | 5 (23.8%) | 11 (47.8%) | | |
| mri_lesional_type: | | | | | | 0.551 | 36 |
| Cavernoma/vascular malformation | 4 (11.1%) | 2 (18.2%) | 0 (0.00%) | 1 (20.0%) | 1 (9.09%) | | |
| Gliosis/stroke | 2 (5.56%) | 0 (0.00%) | 1 (11.1%) | 1 (20.0%) | 0 (0.00%) | | |
| Hippocampal sclerosis | 8 (22.2%) | 4 (36.4%) | 1 (11.1%) | 0 (0.00%) | 3 (27.3%) | | |
| Hippocampal sclerosis + vascular | 2 (5.56%) | 1 (9.09%) | 0 (0.00%) | 0 (0.00%) | 1 (9.09%) | | |
| MCD/FCD | 9 (25.0%) | 1 (9.09%) | 2 (22.2%) | 2 (40.0%) | 4 (36.4%) | | |
| Other | 2 (5.56%) | 0 (0.00%) | 2 (22.2%) | 0 (0.00%) | 0 (0.00%) | | |
| Tumour/glioma | 9 (25.0%) | 3 (27.3%) | 3 (33.3%) | 1 (20.0%) | 2 (18.2%) | | |
